# Network-based Drug Repurposing for Human Coronavirus

**DOI:** 10.1101/2020.02.03.20020263

**Authors:** Yadi Zhou, Yuan Hou, Jiayu Shen, Yin Huang, William Martin, Feixiong Cheng

**Affiliations:** Genomic Medicine Institute, Lerner Research Institute, Cleveland Clinic, Cleveland, OH 44195, USA; Department of Molecular Medicine, Cleveland Clinic Lerner College of Medicine, Case Western Reserve University, Cleveland, OH 44195, USA; Case Comprehensive Cancer Center, Case Western Reserve University School of Medicine, Cleveland, Oh 44106, USA

## Abstract

Human Coronaviruses (HCoVs), including severe acute respiratory syndrome coronavirus (SARS-CoV), Middle east respiratory syndrome coronavirus (MERS-CoV), and 2019 novel coronavirus (2019-nCoV), lead global epidemics with high morbidity and mortality. However, there are currently no effective drugs targeting 2019-nCoV. Drug repurposing, represented as an effective drug discovery strategy from existing drugs, could shorten the time and reduce the cost compared to *de novo* drug discovery. In this study, we present an integrative, antiviral drug repurposing methodology implementing a systems pharmacology-based network medicine platform, quantifying the interplay between the HCoV-host interactome and drug targets in the human protein-protein interaction network. Phylogenetic analyses of 15 HCoV whole genomes reveal that 2019-nCoV has the highest nucleotide sequence identity with SARS-CoV (79.7%) among the six other known pathogenic HCoVs. Specifically, the envelope and nucleocapsid proteins of 2019-nCoV are two evolutionarily conserved regions, having the sequence identities of 96% and 89.6%, respectively, compared to SARS-CoV. Using network proximity analyses of drug targets and known HCoV-host interactions in the human protein-protein interactome, we computationally identified 135 putative repurposable drugs for the potential prevention and treatment of HCoVs. In addition, we prioritized 16 potential anti-HCoV repurposable drugs (including melatonin, mercaptopurine, and sirolimus) that were further validated by enrichment analyses of drug-gene signatures and HCoV-induced transcriptomics data in human cell lines. Finally, we showcased three potential drug combinations (including sirolimus plus dactinomycin, mercaptopurine plus melatonin, and toremifene plus emodin) captured by the ‘*Complementary Exposure’* pattern: the targets of the drugs both hit the HCoV-host subnetwork, but target separate neighborhoods in the human protein-protein interactome network. In summary, this study offers powerful network-based methodologies for rapid identification of candidate repurposable drugs and potential drug combinations toward future clinical trials for HCoVs.

## Introduction

Coronaviruses (CoVs) typically affect the respiratory tract of mammals, including humans, and lead to mild to severe respiratory tract infections ^[1]^. In the past 2 decades, two highly pathogenic human CoVs (HCoVs), including severe acute respiratory syndrome coronavirus (SARS-CoV) and Middle East respiratory syndrome coronavirus (MERS-CoV), emerging from animal reservoirs, have led to global epidemics with high morbidity and mortality^[2]^. For example, 8,098 individuals were infected and 774 died in the SARS-CoV pandemic, which cost the global economy with an estimated $30 to $100 billion^[3, 4]^. According to the World Health Organization (WHO), as of November 2019, MERS-CoV has had a total of 2,494 diagnosed cases causing 858 deaths, the majority in Saudi Arabia^[2]^. In December 2019, the third pathogenic HCoV, named 2019 novel coronavirus (2019-nCoV), was found in Wuhan, China. As of February 02, 2020, there have been over 14,000 cases with ∼300 deaths for the 2019-nCoV pandemic (https://www.cdc.gov/coronavirus/2019-ncov/index.html); furthermore, human-to-human transmission has occurred among close contacts^[5]^. However, there are currently no effective medications against 2019-nCoV. Several national and international research groups are working on the development of vaccines to prevent and treat the 2019-nCoV, but effective vaccines are not available yet. There is an urgent need for the development of effective prevention and treatment strategies for 2019-nCoV outbreak.

Although investment in biomedical and pharmaceutical research and development has increased significantly over the past two decades, the annual number of new treatments approved by the U.S. Food and Drug Administration (FDA) has remained relatively constant and limited^[6]^. A recent study estimated that pharmaceutical companies spent $2.6 billion in 2015, up from $802 million in 2003, in the development of an FDA-approved new chemical entity drug^[7]^. Drug repurposing, represented as an effective drug discovery strategy from existing drugs, could significantly shorten the time and reduce the cost compared to *de novo* drug discovery and randomized clinical trials^[8-10]^. However, experimental approaches for drug repurposing is costly and time-consuming.^[11]^ Computational approaches offer novel testable hypotheses for systematic drug repositioning^[8-10, 12, 13]^. However, traditional structure-based methods are limited when three-dimensional (3D) structures of proteins are unavailable, which, unfortunately, is the case for the majority of human and viral targets. In addition, targeting single virus proteins often have high risk of drug resistance by the rapid evolution of virus genomes ^[1]^.

Viruses (including HCoV) require host cellular factors for successful replication during infections^[1]^. Systematic identification of virus-host protein-protein interactions (PPIs) offers an effective way toward elucidation of the mechanisms of viral infection^[14, 15]^. Subsequently, targeting cellular antiviral targets, such as virus-host interactome, may offer a novel strategy for the development of effective treatments for viral infections^[1]^, including SARS-CoV^[16]^, MERS-CoV^[16]^, Ebola virus^[17]^, and Zika virus^[13, 18-20]^. We recently presented an integrated antiviral drug discovery pipeline that incorporated gene-trap insertional mutagenesis, known functional drug-gene network, and bioinformatics analyses^[13]^. This methodology allows to identify several candidate repurposable drugs for Ebola virus^[10, 13]^. Our work over the last decade has demonstrated how network strategies can, for example, be used to identify effective repurposable drugs^[12, 21-24]^ and drug combinations^[25]^ for multiple human diseases. For example, network-based drug-disease proximity that sheds light on the relationship between drugs (e.g., drug targets) and disease modules (molecular determinants in disease pathobiology modules within the PPIs), and can serve as a useful tool for efficient screening of potentially new indications for approved drugs, as well as drug combinations, as demonstrated in our recent studies^[12, 22, 25]^.

In this study, we present an integrative, antiviral drug repurposing methodology that combines a systems pharmacology-based network medicine platform that quantifies the interplay between the virus-host interactome and drug targets in the human PPI network. The basis for these experiments rests on the notions that (i) the proteins that functionally associate with viral infection (including HCoV) are localized in the corresponding subnetwork within the comprehensive human PPI network^[26]^; and (ii) proteins that serve as drug targets for a specific disease may also be suitable drug targets for potential antiviral infection owing to common PPIs and functional pathways elucidated by the human interactome (**Figure 1**). We follow this analysis with bioinformatics validation of drug-induced gene signatures and HCoV-induced transcriptomics in human cell lines to inspect the postulated mechanism-of-action in a specific HCoV for which we propose repurposing (**Figure 1**).

**Figure 1.**
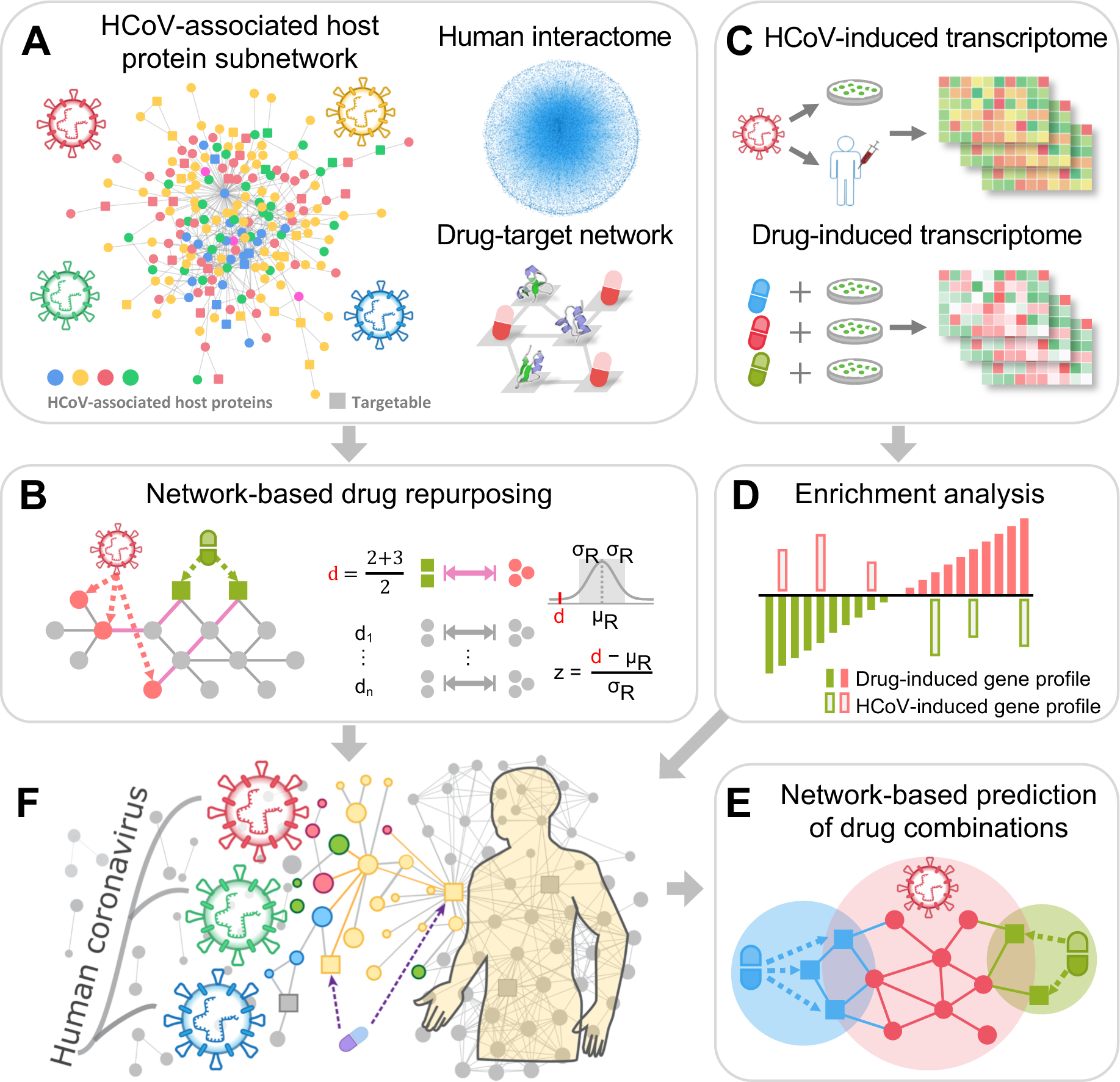
Overall workflow of this study. Our network-based methodology combines a systems pharmacology-based network medicine platform that quantifies the interplay between the virus-host interactome and drug targets in the human PPI network. (**A**) Human coronavirus (HCoV) associated host proteins were collected from literatures and pooled to generate a pan-HCoV protein subnetwork. (**B**) Network proximity between drug targets and HCoV-associated proteins was calculated to screen for candidate repurposable drugs for HCoVs under the human protein interactome model. (**C** & **D**) Gene set enrichment analysis was utilized to validate the network-based prediction. (**E**) Top candidates were further prioritized for drug combinations using network-based method captured by the ‘*Complementary Exposure’* pattern: the targets of the drugs both hit the HCoV-host subnetwork, but target separate neighborhoods in the human interactome network. (**F**) Overall hypothesis of network-based methodology: (i) the proteins that functionally associate with HCoVs are localized in the corresponding subnetwork within the comprehensive human interactome network; and (ii) proteins that serve as drug targets for a specific disease may also be suitable drug targets for potential antiviral infection owing to common protein-protein interactions elucidated by the human interactome.

## Results

### Phylogenetic Analyses of 2019-nCoV

To date, 7 pathogenic HCoVs (**Figure 2A** and **2B**) have been found^[1, 27]^: (i) 2019-nCoV, SARS-CoV, MERS-CoV, HCoV-OC43, and HCoV-HKU1 are β genera, and (ii) HCoV-NL63 and HCoV-229E are α genera. We performed the phylogenetic analyses using the whole genome sequence data from 15 HCoVs to inspect the evolutionary relationship of 2019-nCoV with other HCoVs. We found that the whole genomes of 2019-nCoV had ∼99.99% nucleotide sequence identity across three diagnosed patients (Supplementary **Table S1**). The 2019-nCoV shares the highest nucleotide sequence identity (79.7%) with SARS-CoV among the 6 other known pathogenic HCoVs, revealing conserved evolutionary relationship between 2019-nCoV and SARS-CoV (**Figure 2A**).

**Figure 2.**
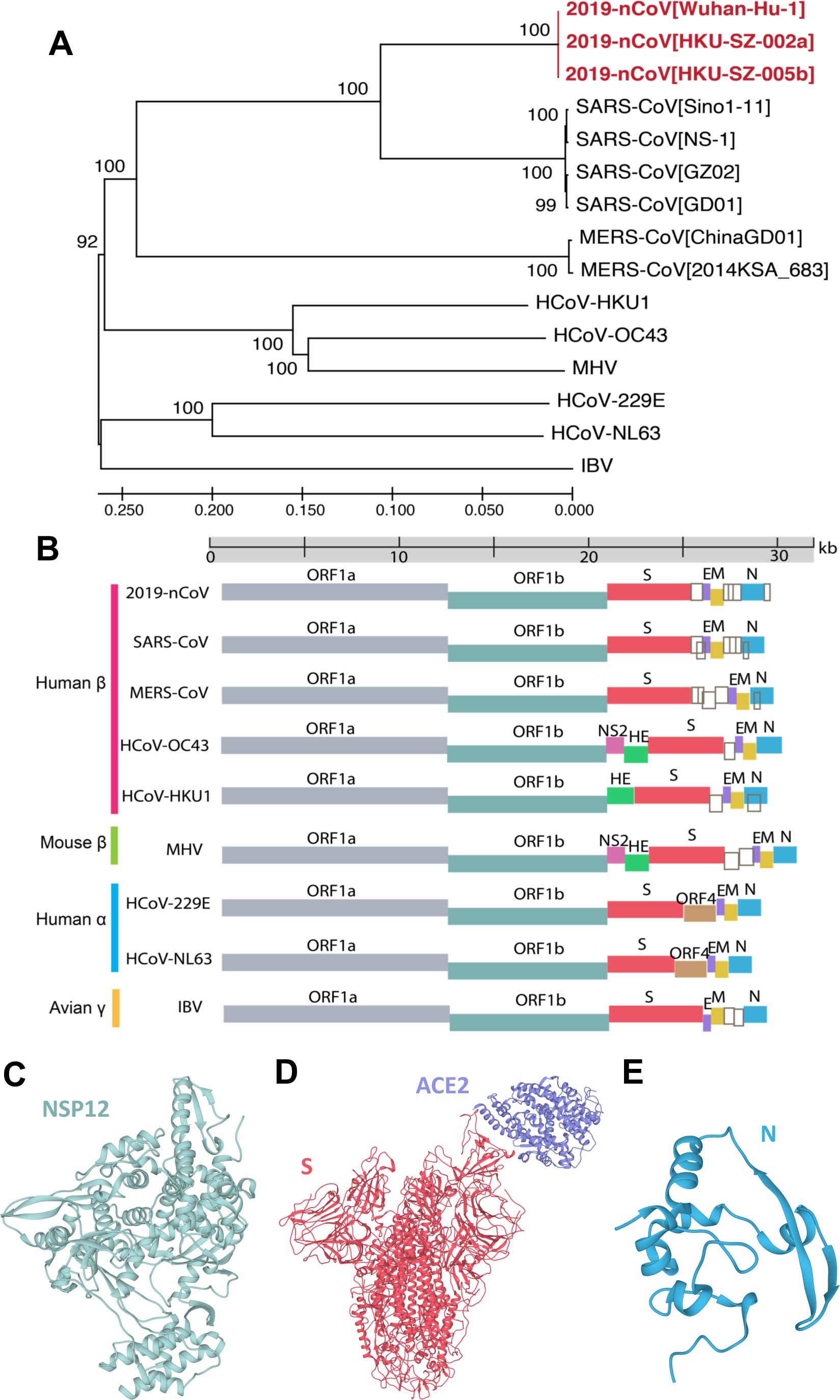
Phylogenetic analysis of coronaviruses. (**A**) Phylogenetic tree of coronavirus (CoV). Phylogenetic algorithm analyzed evolutionary conservation among whole genomes of 15 coronaviruses. Red color highlights the recent emergent coronavirus, 2019-nCoV, originating in Wuhan, China. Numbers on the branches indicate bootstrap support values. The scale shows the evolutionary distance computed using the p-distance method. (**B**) Schematic plot for HCoV genomes. The genus and host information of viruses was labeled on the left by different colors. Empty dark gray boxes represent accessory open reading frames (ORFs). The 3D structures of SARS-CoV nsp12 (PDB ID: 6NUR) (**C**), spike (PDB ID: 6ACK) (**D**) and nucleocapsid (PDB ID: 2CJR) (**E**) shown were based on homology modeling. Genome information and phylogenetic analysis results are provided in Supplementary **Table S1-2**.

HCoVs have five major protein regions for virus structure assembly and viral replications^[27]^, including replicase complex (ORF1ab), spike (S), envelope (E), membrane (M), and nucleocapsid (N) proteins (**Figure 2B**). The ORF1ab gene encodes the non-structural proteins (nsp) of viral RNA synthesis complex through proteolytic processing^[28]^. The nsp12 is a viral RNA-dependent RNA polymerase, together with co-factors nsp7 and nsp8 possessing high polymerase activity. From the protein three-dimensional (3D) structure view of SARS-CoV nsp12, it contains a larger N-terminal extension (which binds to nsp7 and nsp8) and polymerase domain (**Figure 2C**). The spike is a transmembrane glycoprotein that plays a pivotal role in mediating viral infection through binding the host receptor^[29, 30]^. **Figure 2D** shows the 3D structure of the spike protein bound with the host receptor angiotensin converting enznyme2 (ACE2) in SARS-CoV (PDB ID: 6ACK). A recent study showed that 2019-nCoV is able to utilize ACE2 as an entry receptor in ACE2-expressing cells^[31]^, suggesting potential drug targets for therapeutic development. In addition, the nucleocapsid is also an important subunit for packaging the viral genome through protein oligomerization^[32]^, and the single nucleocapsid structure was shown in **Figure 2E**.

Protein sequence alignment analyses indicated that the 2019-nCoV was most evolutionarily conserved with SARS-CoV (Supplementary **Table S2**). Specifically, the envelope and nucleocapsid proteins of 2019-nCoV are two evolutionarily conserved regions, with sequence identities of 96% and 89.6%, respectively, compared to SARS-CoV (Supplementary **Table S2**). However, the spike protein exhibited the lowest sequence conservation (sequence identity of 77%) between 2019-nCoV and SARS-CoV. Meanwhile, the spike protein of 2019-nCoV only has 31.9% sequence identity compared to MERS-CoV.

### HCoV-host Interactome Network

First, we assembled the CoV-associated host proteins from 4 known HCoVs (SARS-CoV, MERS-CoV, HCoV-229E, and HCoV-NL63), one mouse MHV, and one avian IBV (N protein) (Supplementary **Table S3)**. In total, we obtained 119 host proteins associated with CoVs with various experimental evidences. Specifically, these host proteins are either the direct targets of HCoV proteins or are involved in crucial pathways of HCoV infection. The HCoV-host interactome network is shown in **Figure 3A**. We identified several hub proteins including JUN, XPO1, NPM1, and HNRNPA1, with the highest number of connections within the 119 proteins. KEGG pathway enrichment analysis revealed multiple significant biological pathways (adjusted *P* value < 0.05), including measles, RNA transport, NF-kappa B signaling, Epstein-Barr virus infection, and influenza (**Figure 3B**). Gene ontology (GO) biological process enrichment analyses further confirmed multiple viral infection-related processes (adjusted *P* value < 0.001), including viral life cycle, modulation by virus of host morphology or physiology, viral process, positive regulation of viral life cycle, transport of virus, and virion attachment to host cell (**Figure 3C**). We then mapped the known drug-target network (see **Methods**) into the HCoV-host interactome to search for druggable, cellular targets. We found that 47 human proteins (39%, blue nodes in **Figure 3A**) can be targeted by at least one approved drug or experimental drug under clinical trial. For example, GSK3B, DPP4, SMAD3, PARP1, and IKBKB are the most targetable proteins. The high druggability of HCoV-host interactome motivates us to develop a therapeutic strategy by specifically targeting cellular proteins associated with HCoVs, such as drug repurposing.

**Figure 3.**
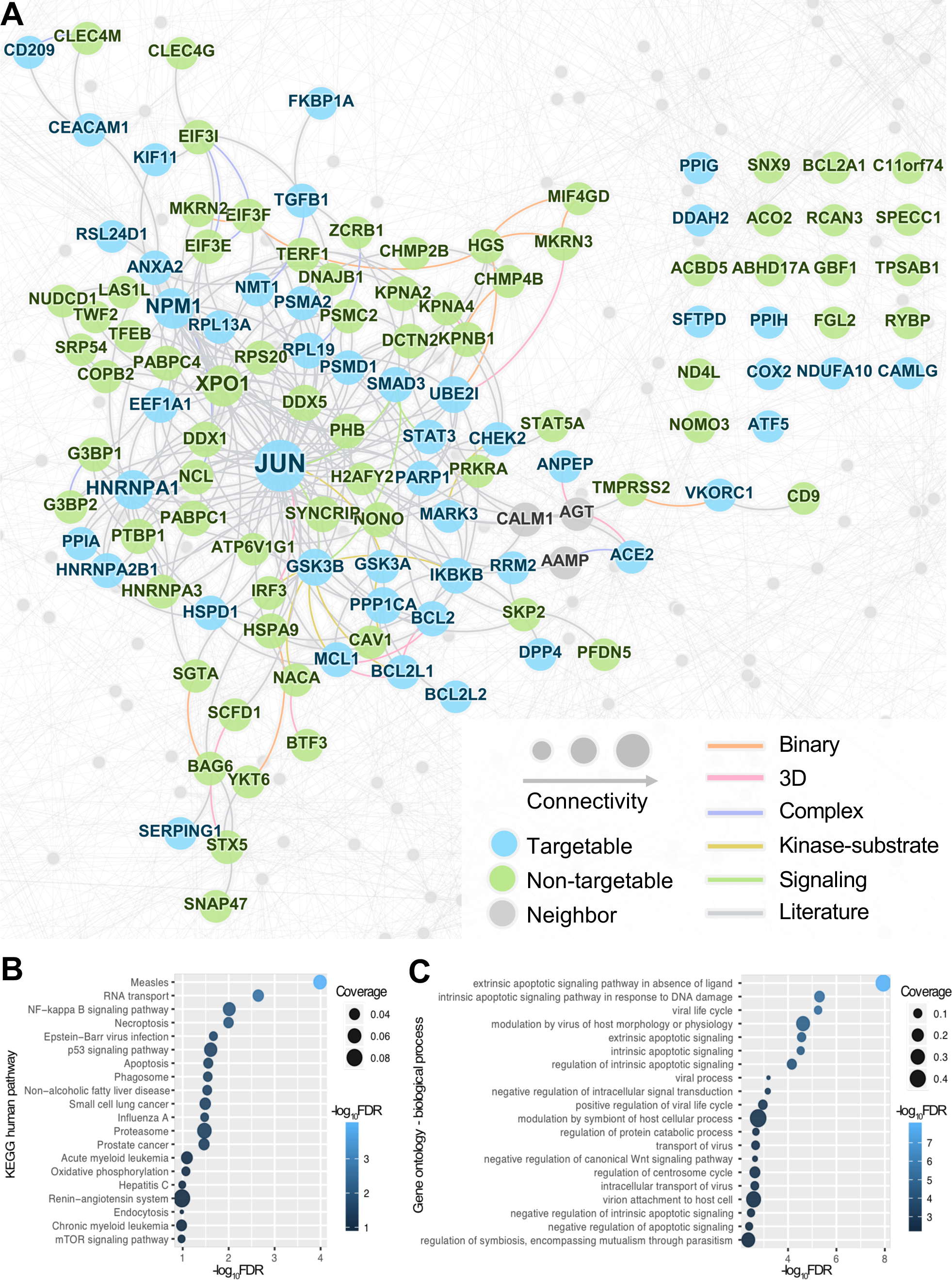
Drug-target network analysis of the HCoV-host interactome. (**A**) A subnetwork highlighting the HCoV-host interactome. Nodes represent three types of HCoV-associated host proteins: targetgable (proteins can be targeted by approved drugs or drugs under clinical trials), non-targetable (proteins don’t have any known ligands), neighbors (protein-protein interaction partners). Edge colors indicate five types of experimental evidences of the protein-protein interactions (see Methods). 3D: three-dimensional structure. (**B**) KEGG human pathway and (**C**) gene ontology enrichment analyses for the HCoV-associated proteins.

### Network-based Drug Repurposing for HCoVs

The basis for the proposed network-based drug repurposing methodologies rests on the notions that the proteins that associate with and functionally govern a viral infection are localized in the corresponding subnetwork (**Figure 1A**) within the comprehensive human interactome network. For a drug with multiple targets to be effective against an HCoV, its target proteins should be within or in the immediate vicinity of the corresponding subnetwork in the human interactome (**Figure 1**), as we demonstrated in multiple diseases ^[12, 21, 22, 25]^ using this drug repurposing strategy. We used a state-of-the-art network proximity measure to quantify the relationship between HCoV-specific subnetwork (**Figure 3A**) and drug targets in the human protein-protein interactome. We constructed a drug-target network by assembling target information for more than 2,000 FDA-approved or experimental drugs (see **Methods**). To improve the quality and completeness of the human protein interactome network, we integrated PPIs with five types of experimental data: (1) binary PPIs from 3D protein structures; (2) binary PPIs from unbiased high-throughput yeast-two-hybrid assays; (3) experimentally identified kinase-substrate interactions; (4) signaling networks derived from experimental data; and (5) literature-derived PPIs with various experimental evidences (see **Methods**). We used a Z-score (Z) measure and permutation test to reduce the study bias in network proximity analyses (including hub nodes in the human interactome network by literature-derived PPI data bias) as described in our recent studies^[12, 25]^.

In total, we computationally identified 135 drugs that were associated (Z < -1.5 and *P* < 0.05, permutation test) with the HCoV-host interactome (**Figure 4A** and Supplementary **Table S4**). To validate bias of the pooled cellular proteins from 6 CoVs, we further calculated the network proximities of all the drugs for 4 CoVs with a high enough number of know host proteins, including SARS-CoV, MERS-CoV, IBV, and MHV, separately. We found that the Z-scores showed consistency among the pooled 119 HCoV-associated proteins and other 4 individual CoVs (**Figure 4B**). The Pearson correlation coefficients of the proximities of all the drugs for the pooled HCoV are 0.926 vs. SARS-CoV (*P* < 0.001, *t* distribution), 0.503 vs. MERS-CoV (*P* < 0.001), 0.694 vs. IBV (*P* < 0.001), and 0.829 vs. MHV (*P* < 0.001). These network proximity analyses offer putative repurposable candidates for potential prevention and treatment of HCoVs.

**Figure 4.**
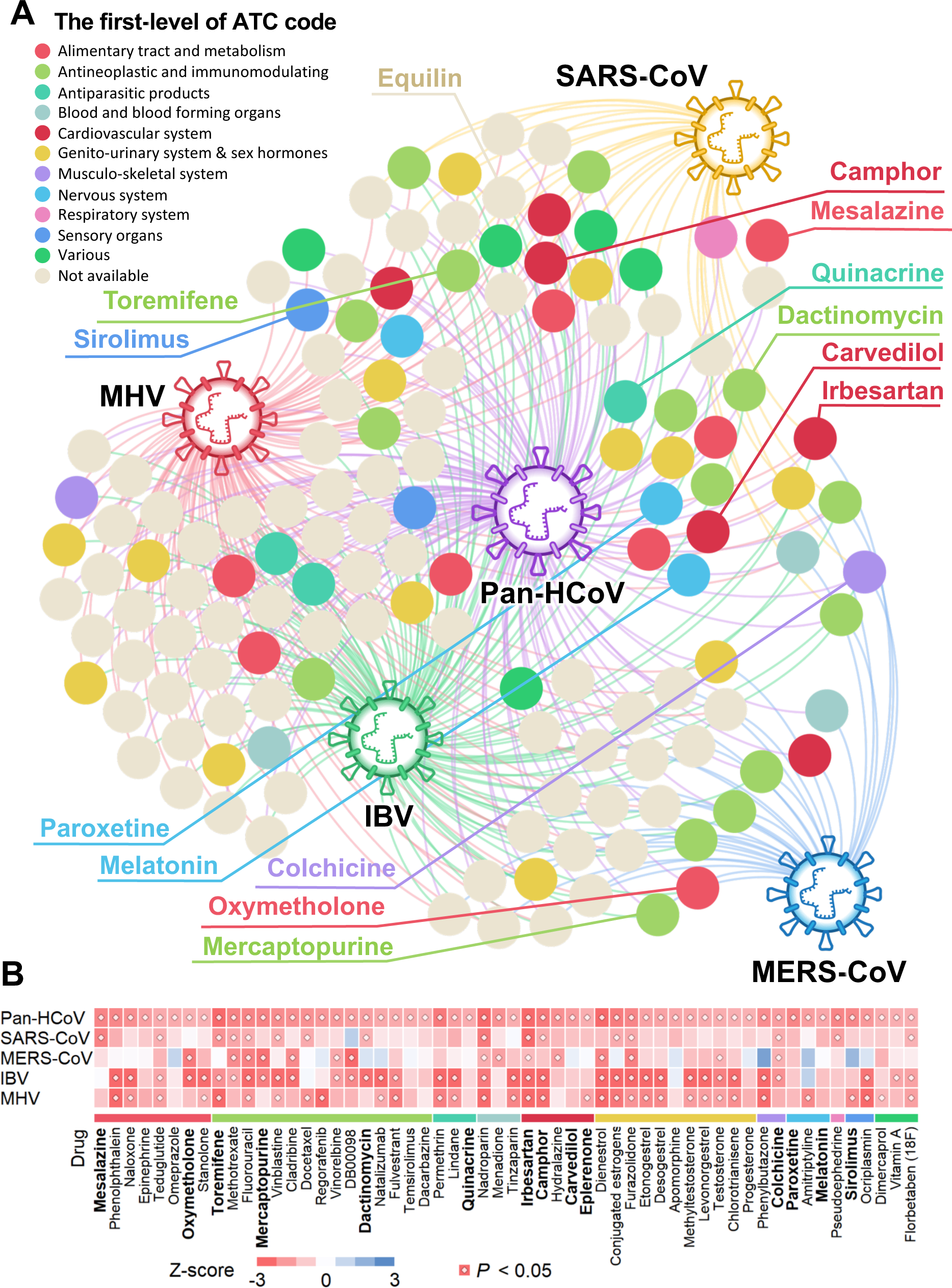
A discovered drug-HCoV network. (**A**) A subnetwork highlighting network-predicted drug-HCoV associations connecting 135 drugs and HCoVs. From the 2,938 drugs evaluated, 135 ones achieved significant proximities between drug targets and the HCoV-associated proteins in the human interactome network. Drugs are colored by their first-level of the Anatomical Therapeutic Chemical (ATC) classification system code. (**B**) A heatmap highlighting network proximity values for SARS-CoV, MERS-CoV, IBV, and MHV respectively. Color key denotes network proximity (Z-score) between drug targets and the HCoV-associated proteins in the human interactome network. P-value was computed by permutation test.

### Discovery of Repurposable Drugs for HCoV

To further validate the 135 repurposable drugs against HCoVs, we first performed gene set enrichment analysis (GSEA) using transcriptome data of MERS-CoV and SARS-CoV infected host cells (see **Methods**). These transcriptome data were used as gene signatures for HCoVs. Additionally, we downloaded the expression data of drug-treated human cell lines from the Connectivity Map (CMAP) database ^[33]^ to obtain drug-gene signatures. We calculated a GSEA score (see **Methods**) for each drug and used this score as an indication of bioinformatics validation of the 135 drugs. Specifically, an enrichment score (*ES*) was calculated for each HCoV data set, and *ES* > 0 and *P* < 0.05 (permutation test) was used as cut-off for a significant association of gene signatures between a drug and a specific HCoV. The GSEA score, ranging from 0 to 3, is the number of data sets that met these criteria for a specific drug. Mesalazine (an approved drug for inflammatory bowel disease), sirolimus (an approved immunosuppressive drug), and equilin (an approved agonist of the estrogen receptor for menopausal symptoms) achieved the highest GSEA scores of 3, followed by paroxetine and melatonin with GSEA scores of 2. We next selected 16 potential repurposable drugs (**Figure 5A** and **Table 1**) against HCoVs using subject matter expertise based on a combination of factors: (i) strength of the network-predicted associations (a smaller network proximity score in Supplementary **Table S4**); (ii) validation by GSEA analyses; (iii) literature-reported antiviral evidences, and (iv) fewer clinically reported side effects. Specifically, we showcased several selected repurposable drugs with literature-reported antiviral evidences as below.

**Table 1.**
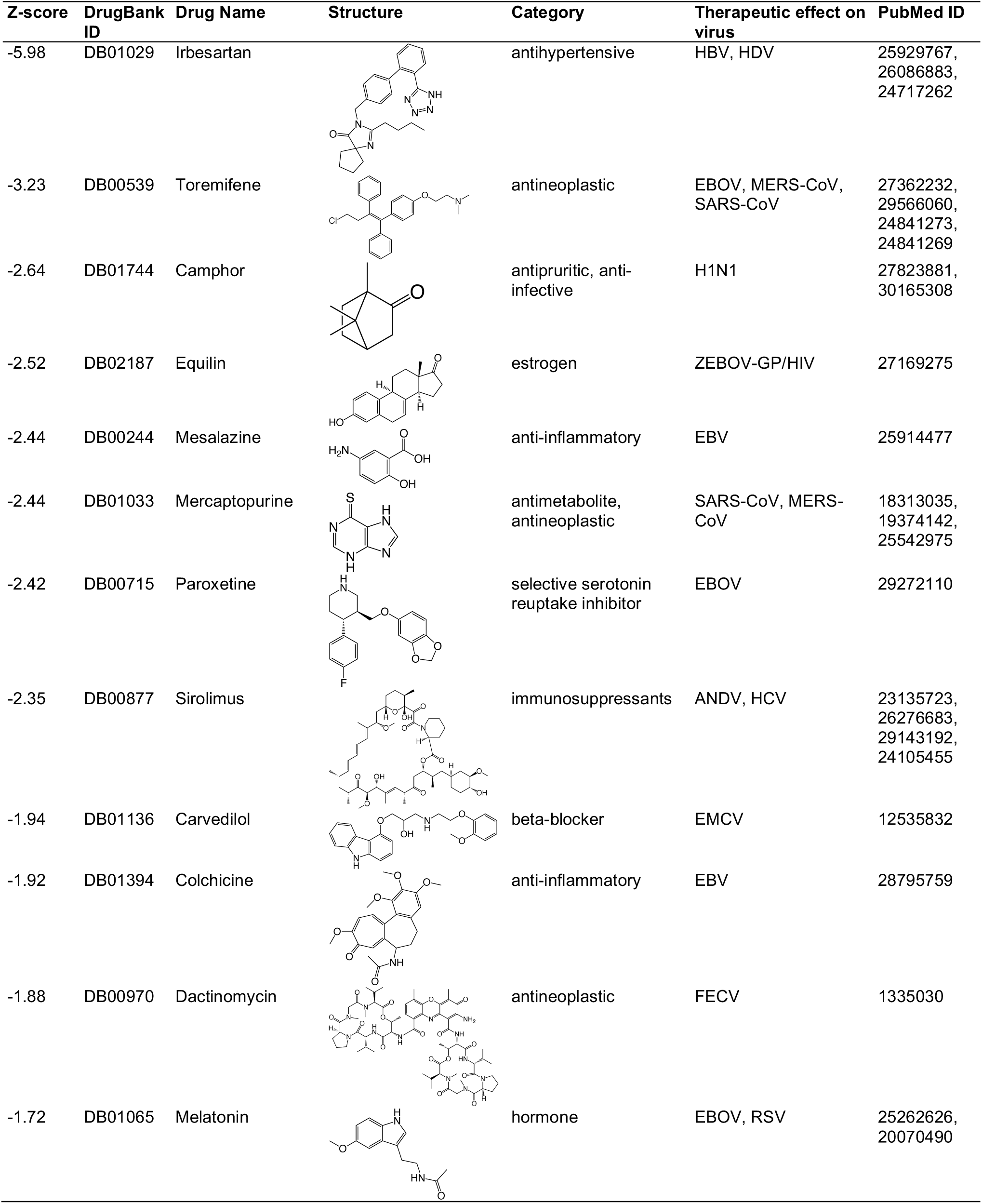
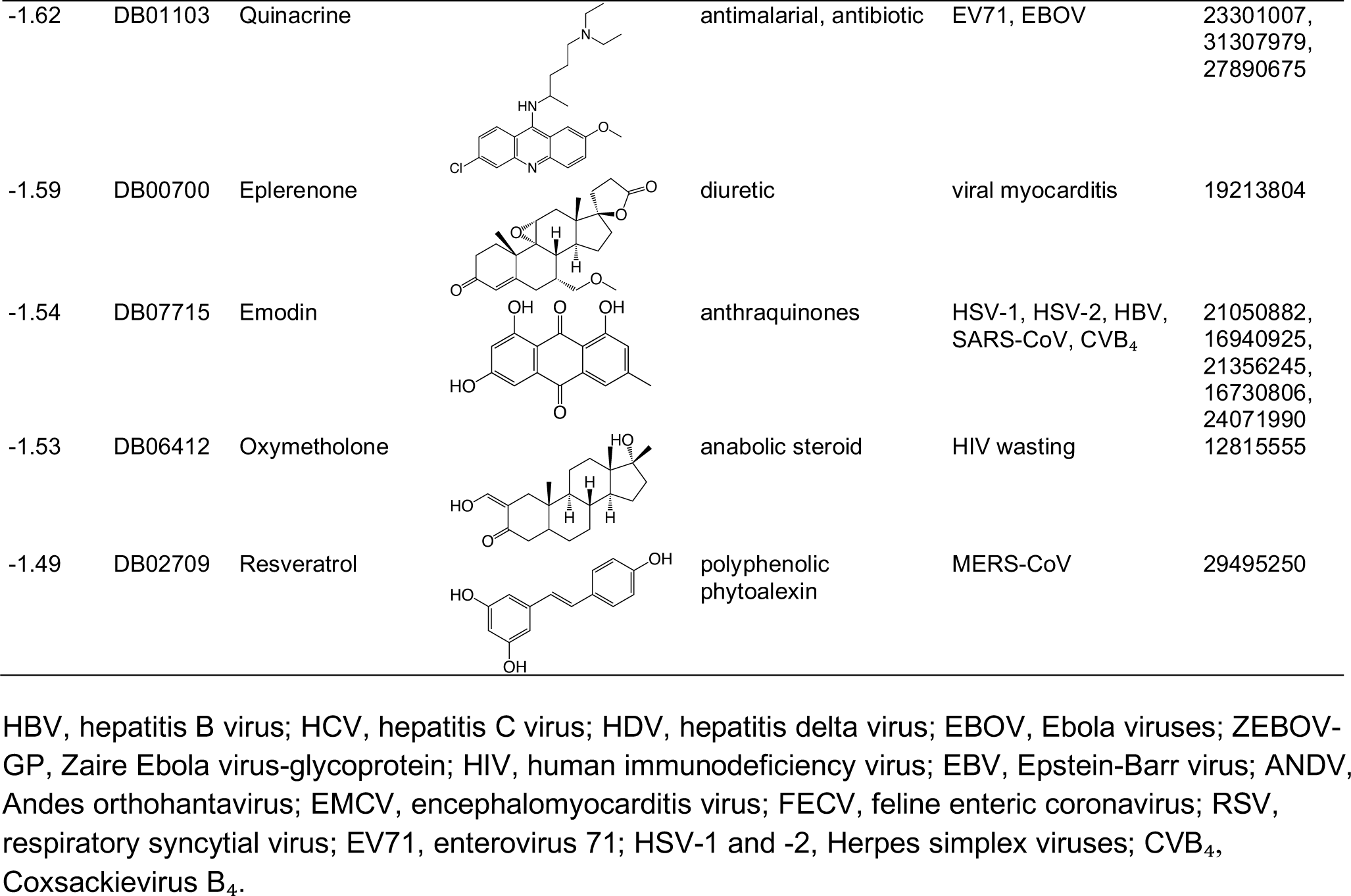
Top drug candidates from network-based repurposing approach with literature antiviral evidences.

| Z-score | DrugBank ID | Drug Name | Structure | Category | Therapeutic effect on virus | PubMed ID |
| --- | --- | --- | --- | --- | --- | --- |
| -5.98 | DB01029 | Irbesartan |  | antihypertensive | HBV, HDV | 25929767,<br>26086883,<br>24717262 |
| -3.23 | DB00539 | Toremifene |  | antineoplastic | EBOV, MERS-CoV, SARS-CoV | 27362232,<br>29566060,<br>24841273,<br>24841269 |
| -2.64 | DB01744 | Camphor |  | antipruritic, anti-infective | H1N1 | 27823881,<br>30165308 |
| -2.52 | DB02187 | Equilin |  | estrogen | ZEBOV-GP/HIV | 27169275 |
| -2.44 | DB00244 | Mesalazine |  | anti-inflammatory | EBV | 25914477 |
| -2.44 | DB01033 | Mercaptopurine |  | antimetabolite, antineoplastic | SARS-CoV, MERS-CoV | 18313035,<br>19374142,<br>25542975 |
| -2.42 | DB00715 | Paroxetine |  | selective serotonin reuptake inhibitor | EBOV | 29272110 |
| -2.35 | DB00877 | Sirolimus |  | immunosuppressants | ANDV, HCV | 23135723,<br>26276683,<br>29143192,<br>24105455 |
| -1.94 | DB01136 | Carvedilol |  | beta-blocker | EMCV | 12535832 |
| -1.92 | DB01394 | Colchicine |  | anti-inflammatory | EBV | 28795759 |
| -1.88 | DB00970 | Dactinomycin |  | antineoplastic | FECV | 1335030 |
| -1.72 | DB01065 | Melatonin |  | hormone | EBOV, RSV | 25262626,<br>20070490 |
| -1.62 | DB01103 | Quinacrine |  | antimalarial, antibiotic | EV71, EBOV | 23301007,<br>31307979,<br>27890675 |
| -1.59 | DB00700 | Eplerenone |  | diuretic | viral myocarditis | 19213804 |
| -1.54 | DB07715 | Emodin |  | anthraquinones | HSV-1, HSV-2, HBV,<br>SARS-CoV, CVB<sub>4</sub> | 21050882,<br>16940925,<br>21356245,<br>16730806,<br>24071990 |
| -1.53 | DB06412 | Oxymetholone |  | anabolic steroid | HIV wasting | 12815555 |
| -1.49 | DB02709 | Resveratrol |  | polyphenolic<br>phytoalexin | MERS-CoV | 29495250 |

HBV, hepatitis B virus; HCV, hepatitis C virus; HDV, hepatitis delta virus; EBOV, Ebola viruses; ZEBOV-GP, Zaire Ebola virus-glycoprotein; HIV, human immunodeficiency virus; EBV, Epstein-Barr virus; ANDV, Andes orthohantavirus; EMCV, encephalomyocarditis virus; FECV, feline enteric coronavirus; RSV, respiratory syncytial virus; EV71, enterovirus 71; HSV-1 and -2, Herpes simplex viruses; CVB<sub>4</sub>, Coxsackievirus B<sub>4</sub>.

**Figure 5.**
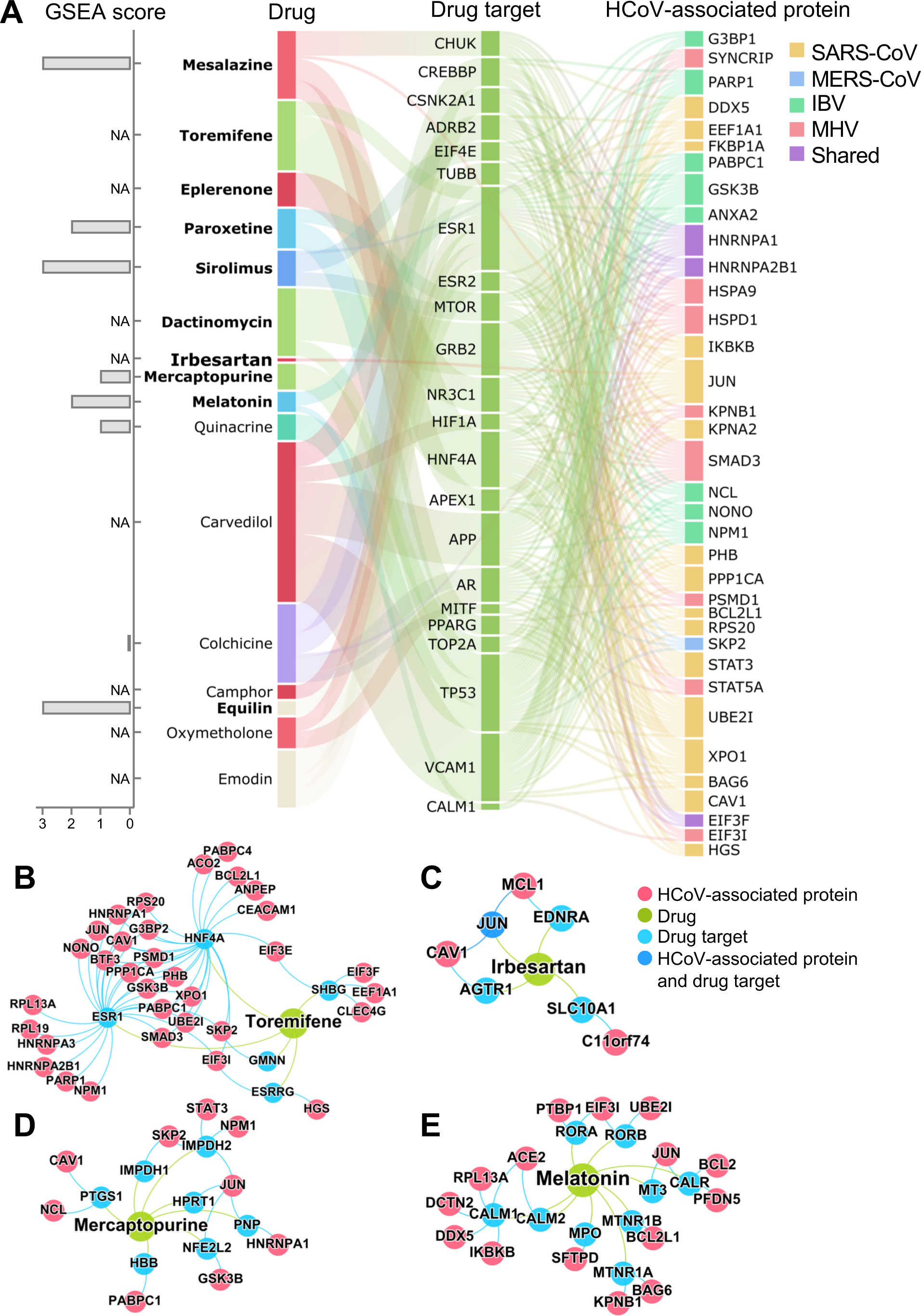
A discovered drug-protein-HCoV network for 16 potential repurposable drugs. (**A**) Network-predicted evidences and gene set enrichment analysis (GSEA) scores for 16 potential repurposable drugs for HCoVs. The overall connectivity of the top drug candidates to the HCoV-associated proteins was examined. Most of these drugs indirectly target HCoV-associated proteins via the human protein-protein interaction networks. All the drug-target-HCoV-associated protein connections were examined, and those proteins with at least 5 connections are shown. The box heights for the proteins indicate the number of connections. GSEA scores for 8 drugs were not available (NA) due to the lack of transcriptome profiles for the drugs. (**B-E**) Inferred mechanism-of-action networks for four selected drugs: (**B**) toremifene (first generation nonsteroidal selective estrogen receptor modulator), (**C**) irbesartan (an angiotensin receptor blocker), (**D**) mercaptopurine (an antimetabolite antineoplastic agent with immunosuppressant properties), and (**E**) melatonin (a biogenic amine for treating circadian rhythm sleep disorders).

#### Selective estrogen receptor modulators (SERMs)

An overexpression of estrogen receptor has been shown to play a crucial role in inhibiting viral replication^[34]^. SERMs have been reported to play a broader role in inhibiting viral replication through the non-classical pathways associated with estrogen receptor^[34]^. SERMs interfere at the post viral entry step and affect the triggering of fusion, as the SERMs’ antiviral activity still can be observed in the absence of detectable estrogen receptor expression^[17]^. Toremifene (Z = -3.23, **Figure 5A**), the first generation of nonsteroidal SERM, exhibits potential effects in blocking various viral infections, including MERS-CoV, SARS-CoV, and Ebola virus in established cell lines^[16, 35]^. Interestingly, different from the classical ESR1-related antiviral pathway, toremifene prevents fusion between the viral and endosomal membrane by interacting with and destabilizing the virus membrane glycoprotein, and eventually inhibiting viral replication^[36]^. As shown in **Figure 5B**, toremifene potentially affects several key host proteins associated with HCoV, such as RPL19, HNRNPA1, NPM1, EIF3I, EIF3F, and EIF3E^[37, 38]^. Equilin (Z = -2.52 and GSEA score = 3), an estrogenic steroid produced by horses, also has been proven to have moderate activity in inhibiting the entry of Zaire Ebola virus-glycoprotein and human immunodeficiency virus (ZEBOV-GP/HIV)^[17]^. Altogether, network-predicted SERMs (such as toremifene and equilin) offer potential repurposable candidates for HCoVs.

#### Angiotensin receptor blockers (ARBs)

ARBs have been reported to associate with viral infection, including HCoVs^[39-41]^. Irbesartan (Z = -5.98), a typical ARB, was approved by the FDA for treatment of hypertension and diabetic nephropathy. Here, network proximity analysis shows a significant association between Irbesartan’s targets and HCoV-associated host proteins in the human interactome. As shown in **Figure 5C**, irbesartan targets SLC10A1, encoding the sodium/bile acid cotransporter (NTCP) protein that has been identified as a functional preS1-specific receptor for the hepatitis B virus (HBV) and the hepatitis delta virus (HDV). Irbesartan can inhibit NTCP, thus inhibiting viral entry^[42, 43]^. SLC10A1 interacts with C11orf74, a potential transcriptional repressor that interacts with nsp-10 of SARS-CoV^[44]^. There are several other ARBs (such as eletriptan, frovatriptan, and zolmitriptan) in which their targets are potentially associated with HCoV-associated host proteins in the human interactome.

#### Immunosuppressant or antineoplastic agents

Previous studies have confirmed the mammalian target of rapamycin complex 1 (mTORC1) as the key factor in regulating various viruses’ replications, including Andes orthohantavirus and coronavirus^[45, 46]^. Sirolimus (Z = -2.35 and GSEA score = 3), an inhibitor of mammalian target of rapamycin (mTOR), was reported to effectively block viral protein expression and virion release effectively^[47]^. Indeed, the latest study revealed the clinical application: sirolimus reduced MERS-CoV infection by over 60%^[48]^. Moreover, sirolimus usage in managing patients with severe H1N1 pneumonia and acute respiratory failure can improve those patients’ prognosis significantly^[47]^. Mercaptopurine (Z = -2.44 and GSEA score = 1), an antineoplastic agent with immunosuppressant property, has been used to treat cancer since the 1950s and expanded its application to several auto-immune diseases, including rheumatoid arthritis, systemic lupus erythematosus, and Crohn’s disease^[49]^. Mercaptopurine has been reported as a selective inhibitor of both SARS-CoV and MERS-CoV by targeting papain-like protease which plays key roles in viral maturation and antagonism to interferon stimulation^[50, 51]^. Mechanistically, mercaptopurine potentially target several host proteins in HCoVs, such as JUN, PABPC1, NPM1, and NCL^[37, 52]^ (**Figure 5D**).

#### Anti-inflammatory agents

Inflammatory pathways play essential roles in viral infections^[53, 54]^. As a biogenic amine, melatonin (N-acetyl-5-methoxytryptamine) (Z = -1.72 and GSEA score = 2) plays a key role in various biological processes, and offers a potential strategy in the management of viral infections^[55, 56]^. Viral infections are often associated with immune-inflammatory injury, in which the level of oxidative stress increases significantly and leaves negative effects on multiple organ functions^[57]^. The antioxidant effect of melatonin makes it a putative candidate drug to relieve patients’ clinical symptoms in antiviral treatment, even though melatonin cannot eradicate or even curb the viral replication or transcription^[58, 59]^. In addition, the application of melatonin may prolong patients’ survival time, which may provide a chance for patients’ immune systems to recover and eventually eradicate the virus. As shown in **Figure 5E**, melatonin indirectly targets several HCoV cellular targets, including ACE2, BCL2L1, JUN, and IKBKB. Eplerenone (Z = -1.59), an aldosterone receptor antagonist, is reported to have a similar anti-inflammatory effect as melatonin. By inhibiting mast-cell-derived proteinases and suppressing fibrosis, eplerenone can improve survival of mice infected with encephalomyocarditis virus^[60]^.

In summary, our network proximity analyses offer multiple putative repurposable drugs that target diverse cellular pathways for potential prevention and treatment of HCoVs. However, further preclinical experiments and clinical trials are required to verify the clinical benefits of these network-predicted candidates before clinical use.

### Network-based Identification of Potential Drug Combinations for HCoV

Drug combinations, offering increased therapeutic efficacy and reduced toxicity, play an important role in treating various viral infections^[61]^. However, our ability to identify and validate effective combinations is limited by a combinatorial explosion, driven by both the large number of drug pairs and dosage combinations. In our recent study, we proposed a novel network-based methodology to identify clinically efficacious drug combinations^[25]^. Relying on approved drug combinations for hypertension and cancer, we found that a drug combination was therapeutically effective only if it was captured by the ‘*Complementary Exposure*’ pattern: the targets of the drugs both hit the disease module, but target separate neighborhoods (**Figure 6A**). Here we sought to identify drug combinations that may provide a synergistic effect in treating HCoVs with well-defined mechanism-of-action by network analysis. For the 16 potential repurposable drugs (**Figure 5A**), we showcased three network-predicted candidate drug combinations in the potential treatment of HCoVs.

**Figure 6.**
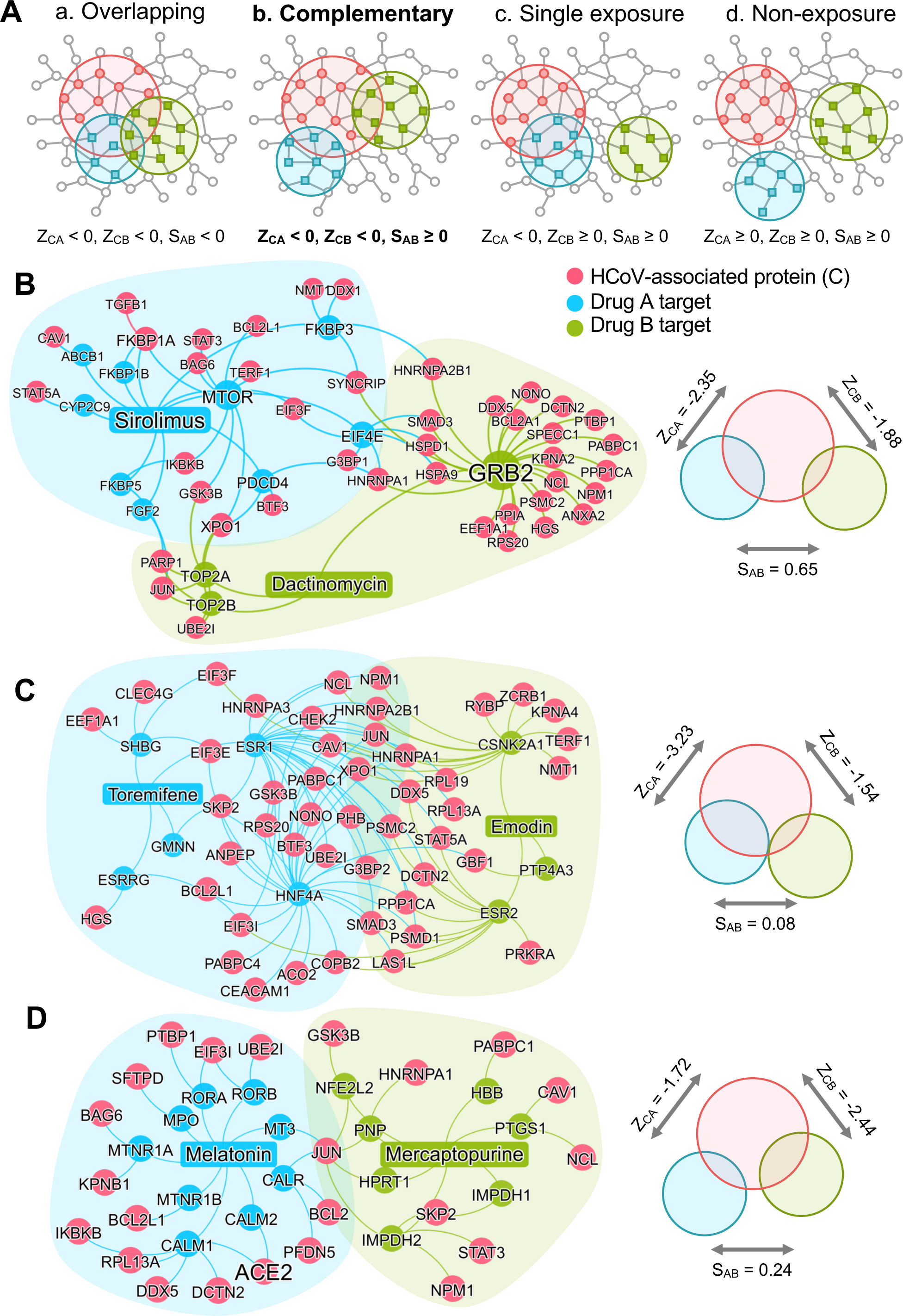
Network-based rational design of drug combinations for HCoVs. (**A**) The possible exposure mode of the HCoV-associated protein module to the pairwise drug combinations. An effective drug combination will be captured by the ‘*Complementary Exposure’* pattern: the targets of the drugs both hit the HCoV-host subnetwork, but target separate neighborhoods in the human interactome network. ZCA and ZCB denote the network proximity (Z-score) between targets (Drugs A and B) and a specific HCoV. S_AB_ denotes separation score (see Methods) of targets between Drug A and Drug B. (**B-D**) Inferred mechanism-of-action networks for three selected pairwise drug combinations: (**B**) Sirolimus (a potent immunosuppressant with both antifungal and antineoplastic properties) plus Dactinomycin (a RNA synthesis inhibitor for treatment of various tumors), (**C**) Toremifene (first generation nonsteroidal selective estrogen receptor modulator) plus Emodin (an experimental drug for the treatment of polycystic kidney), and (**D**) Melatonin (a biogenic amine for treating circadian rhythm sleep disorders) plus Mercaptopurine (an antimetabolite antineoplastic agent with immunosuppressant properties).

#### Sirolimus plus Dactinomycin

Sirolimus, an inhibitor of mTOR with both antifungal and antineoplastic properties, has demonstrated to improves outcomes in patients with severe H1N1 pneumonia and acute respiratory failure^[47]^. The mTOR signaling plays an essential role for MERS-CoV infection^[62]^. Dactinomycin, also known actinomycin D, is an approved RNA synthesis inhibitor for treatment of various cancer types. An early study showed that dactinomycin (1 microgram/ml) inhibited the growth of feline enteric CoV^[63]^. As shown in **Figure 6B**, our network analysis shows that sirolimus and dactinomycin synergistically target HCoV-associated host protein subnetwork by ‘Complementary Exposure’ pattern, offering potential combination regimens for treatment of HCoV. Specifically, sirolimus and dactinomycin may inhibit both mTOR signaling and RNA synthesis pathway (including DNA topoisomerase 2-alpha (TOP2A) and DNA topoisomerase 2-beta (TOP2B)) in HCoV infected cells (**Figure 6B**).

#### Toremifene plus Emodin

Toremifene is the approved first generation nonsteroidal SERMs for the treatment of metastatic breast cancer^[64]^. SERMs (including toremifene) inhibited Ebola virus infection^[17]^ by interacting with and destabilizing the Ebola virus glycoprotein^[36]^. *In vitro* assays have demonstrated that toremifene inhibited growth of MERS-CoV^[16, 65]^ and SARA-CoV^[35]^ (**Table 1**). Emodin, an anthraquinone derivative extracted from the roots of rheum tanguticum, have been reported to have various anti-virus effects. Specifically, emdoin inhibited SARS-CoV associated 3a protein^[66]^, and blocked an interaction between the SARS-CoV spike protein and ACE2^[67]^. Altogether, network analyses and published experimental data suggested that combining toremifene and emdoin offered a potential therapeutic approach for HCoVs (**Figure 6C**).

#### Mercaptopurine plus Melatonin

As shown in **Figure 5A**, targets of both mercaptopurine and melatonin showed strong network proximity with HCoV-associated host proteins in the human interactome network. Recent *in vitro* and *in vivo* studies identified mercaptopurine as a selective inhibitor of both SARS-CoV and MERS-CoV by targeting papain-like protease ^[50, 51]^. Melatonin was reported in potential treatment of viral infection via its anti-inflammatory and antioxidant effects^[55-59]^. Melatonin indirectly regulates ACE2 expression, a key entry receptor involved in viral infection of HCoVs, including 2019-nCoV^[31]^. JUN, also known as c-Jun, is a key host protein involving in HCoV infectious bronchitis virus^[68]^. As shown in **Figure 6D**, mercaptopurine and melatonin may synergistically block c-Jun signaling by targeting multiple cellular targets. In summary, combination of mercaptopurine and melatonin may offer a potential combination therapy for 2019-nCoV by synergistically targeting papain-like protease, ACE2, c-Jun signaling, and anti-inflammatory pathways (**Figure 6D**). However, further experimental and clinical validations are highly warranted.

## Discussion

In this study, we presented a network-based methodology for systematic identification of putative repurposable drugs and drug combinations for potential treatment of HCoV. Integration of drug-target networks, HCoV-host interactions, HCoV-induced transcriptome in human cell lines, and human protein-protein interactome network are essential for such identification. Based on comprehensive evaluation, we prioritized 16 putative repurposable drugs (**Figure 5**) and 3 putative drug combinations (**Figure 6**) for the potential treatment of HCoVs, including 2019-nCoV. However, all network-predicted repurposable drugs and drug combinations must be validated in preclinical models and randomized clinical trials before being used in patients.

We acknowledge several limitations in our current study. In this study, we used a low binding affinity value of 10 μM as a threshold to define a physical drug-target interaction. However, a stronger binding affinity threshold (e.g., 1μM) may be a more suitable cut-off in drug discovery, although it will generate a smaller drug-target network. Although sizeable efforts were made for assembling large-scale, experimentally reported drug-target networks from publicly available databases, the network data may be incomplete and some drug-protein interactions may be functional associations, instead of physical bindings. We may use computational approaches to systematically predict the drug-target interactions further^[24, 69]^. In addition, the collected virus-host interactions are far from complete and the quality can be influenced by multiple factors, including different experimental assays and human cell line models. We may computationally predict a new virus-host interactome for HCoVs using sequence-based and structure-based approaches^[70]^. The current systems pharmacology model cannot separate therapeutic antiviral effects from those predictions due to lack of detailed pharmacological effects of drug targets and unknown functional consequences of virus-host interactions. Drug targets representing nodes within cellular networks are often intrinsically coupled with both therapeutic and adverse profiles^[71]^, as drugs can inhibit or activate protein functions (including antagonists versus agonists). Comprehensive identification of the virus-host interactome for 2019-nCoV, with specific biological effects using functional genomics assays^[72, 73]^, will significantly improve the accuracy of current network-based methodologies.

Owing to lack of the complete drug target information (such as the molecular ‘promiscuity’ of drugs), the dose-response and dose-toxicity effects for both repurposable drugs and drug combinations cannot be identified in current network models. For example, Mesalazine, an approved drug for inflammatory bowel disease, is a top network-predicted candidate drug (**Figure 5A**) associated with HCoVs. Yet, several clinical studies showed the potential pulmonary toxicities (including pneumonia) associated with mesalazine usage ^[74, 75]^. Preclinical studies are warranted to evaluate *in vivo* efficiency and side effects before clinical trials. Furthermore, we only limited to predict pairwise drug combinations based on our previous network-based framework^[25]^. However, we expect that our methodology reminds to be a useful network-based tools for prediction of combining multiple drugs toward exploring network relationships of multiple drugs’ targets with the HCoV-host subnetwork in the human interactome. Finally, we aimed to systematically identify repurposable drugs by specifically targeting nCoV host proteins only. Thus, our current network models cannot predict repurposable drugs from the existing anti-virus drugs that target virus proteins only. Thus, combination of the existing anti-virus drugs (such as remdesivir^[76]^) with the network-predicted repurposable drugs (**Figure 5**) or drug combinations (**Figure 6**) may improve coverage of current network-based methodologies by utilizing multi-layer network framework.

In conclusion, this study offers a powerful, integrated network-based systems pharmacology methodology for rapid identification of repurposable drugs and drug combinations for the potential treatment of HCoV. Our approach can minimize the translational gap between preclinical testing results and clinical outcomes, which is a significant problem in the rapid development of efficient treatment strategies for the emerging 2019-nCoV outbreak. From a translational perspective, if broadly applied, the network tools developed here could help develop effective treatment strategies for other types of virus and human diseases as well.

## Methods and Materials

### Genome Information and Phylogenetic analysis

In total, we collected DNA sequences and protein sequences for 15 HCoVs, including three most recent 2019-nCoV genomes, from the NCBI GenBank database (January 28, 2019, Supplementary **Table S1**). Whole genome alignment and protein sequence identity calculation were performed by Multiple Sequence Alignment in EMBL-EBI database with default parameters. The neighbor joining (NJ) tree was computed from the pairwise phylogenetic distance matrix using MEGA X^[77]^ with 1000 bootstrap replicates. The protein alignment and phylogenetic tree of HCoVs were constructed by MEGA X.

### Building the Virus-Host Interactome

We collected HCoV-host protein interactions from various literatures based on our sizeable efforts. The HCoV-associated host proteins of several HCoVs, including SARS-CoV, MERS-CoV, IBV, MHV, HCoV-229E, and HCoV-NL63 were pooled. These proteins were either the direct targets of HCoV proteins or were involved in critical pathways of HCoV infection identified by multiple experimental sources, including high throughput yeast-two-hybrid (Y2H) systems, viral protein pull-down assay, *in vitro* co-immunoprecipitation and RNA knock down experiment. In total, the virus-host interaction network included 6 HCoVs with 119 host proteins (Supplementary **Table S3**).

### Functional Enrichment Analysis

Next, we performed Kyoto Encyclopedia of Genes and Genomes (KEGG) and Gene Ontology (GO) enrichment analyses to evaluate the biological relevance and functional pathways of the HCoV-associated proteins. All functional analyses were performed using Enrichr^[78]^.

### Building the Drug-Target Network

Here, we collected drug-target interaction information from the DrugBank database (v4.3)^[79]^, Therapeutic Target Database (TTD)^[80]^, PharmGKB database, ChEMBL (v20)^[81]^, BindingDB^[82]^, and IUPHAR/BPS Guide to PHARMACOLOGY^[83]^. The chemical structure of each drug with SMILES format was extracted from DrugBank^[79]^. Here, only drug-target interactions meeting the following three criteria were used: (i) binding affinities, including Ki, Kd, IC50 or EC_50_ each ≤ 10 μM; (ii) the target was marked as ‘reviewed’ in the UniProt database^[84]^; and (iii) the human target was represented by a unique UniProt accession number. The details for building the experimentally validated drug-target network are provided in our recent study^[12]^.

### Building the Human Protein-Protein Interactome

To build a comprehensive list of human PPIs, we assembled data from a total of 18 bioinformatics and systems biology databases with five types of experimental evidences: (i) Binary PPIs tested by high-throughput yeast-two-hybrid (Y2H) systems (ii) Binary, physical PPIs from protein three-dimensional (3D) structures; (iii) Kinase-substrate interactions by literature-derived low-throughput or high-throughput experiments; (iv) Signaling network by literature-derived low-throughput experiments; and (v) Literature-curated PPIs identified by affinity purification followed by mass spectrometry (AP-MS), Y2H, or by literature-derived low-throughput experiments. All inferred data, including evolutionary analysis, gene expression data, and metabolic associations, were excluded. The genes were mapped to their Entrez ID based on the NCBI database^[85]^ as well as their official gene symbols based on GeneCards (https://www.genecards.org/). In total, the resulting human protein-protein interactome used in this study includes 351,444 unique PPIs (edges or links) connecting 17,706 proteins (nodes), representing a 50% increase in the number of the PPIs we have used previously. Detailed descriptions for building the human protein-protein interactome are provided in our previous studies^[12, 86, 87]^.

### Network Proximity Measure

We posit that the human PPIs provide an unbiased, rational roadmap for repurposing drugs for potential treatment of HCoVs in which they were not originally approved. Given *c* the set of host genes associated with a specific HCoV, and *T* the set of drug targets, we computed the network proximity of *C* with the target set *T* of each drug using the “closest” method:

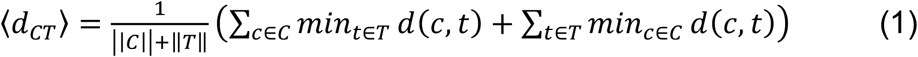

where *d*(*c, t*) is the shortest distance between gene *c* and *t* in the human protein interactome. The network proximity was converted to Z-score based on permutation tests:

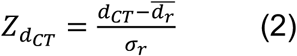

where 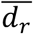 and *σ*_r_ were the mean and standard deviation of the permutation test repeated 1,000 times, each time with two randomly selected gene lists with similar degree distributions to those of *C* and *T*. The corresponding *P* value was calculated based on the permutation test results. Z-score < -1.5 and *P* < 0.05 were considered significantly proximal drug-HCoV associations. All networks were visualized using Gephi 0.9.2 (https://gephi.org/).

### Network-based Rational Design of Drug Combination Prediction

For this network-based approach for drug combinations to be effective, we need to establish if the topological relationship between two drug-target modules reflects biological and pharmacological relationships, while also quantifying their network-based relationship between drug-targets and HCoV-associated host proteins (drug-drug-HCoV combinations). To identify potential drug combinations, we combined the top lists of drugs. Then, “separation” measure *S*_*AB*_ was calculated for each pair of drugs *A* and *B* using the following method:

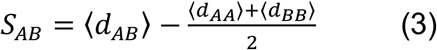

where ⟨*d*.⟩ was calculated based on the “closest” method. Our key methodology is that a drug combination is therapeutically effective only if it follows a specific relationship to the disease module, as captured by *Complementary Exposure* patterns in targets’ modules of both drugs without overlapping toxic mechanisms.^[25]^

### Gene Set Enrichment Analysis

We performed the gene set enrichment analysis as an additional prioritization method. We first collected three differential gene expression data sets of hosts infected by HCoVs from the NCBI Gene Expression Omnibus (GEO). Among them, two transcriptome data sets were SARS-CoV infected samples from patient’s peripheral blood^[88]^ (GSE1739) and Calu-3 cells^[89]^ (GSE33267) respectively. One transcriptome data set was MERS-CoV infected Calu-3 cells^[90]^ (GSE122876). Adjusted *P* value less than 0.01 was defined as differentially expressed genes. These data sets were used as HCoV host signatures to evaluate the treatment effects of drugs. Differential gene expression in cells treated with various drugs were retrieved from the Connectivity Map (CMAP) database^[33]^, and were used as gene profiles for the drugs. For each drug that was in both the CMAP data set and our drug-target network, we calculated an enrichment score (*ES*) for each HCoV signature data set based on previously described methods^[91]^ as follows:

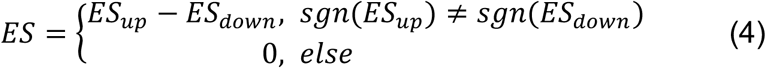

*ES*_*up*_and *ES*_*down*_ were calculated separately for the up- and down-regulated genes from the HCoV signature data set using the same method. We first computed *a*_*up/down*_ and *b*_*up/down*_ as:

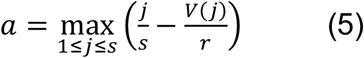

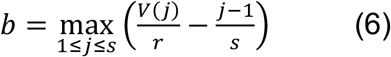

Where *j=1,2, …,s* were the genes of HCoV signature data set sorted in ascending order by their rank in the gene profiles of the drug being evaluated. The rank of gene *j* is denoted by *V*(*j*), where 1 ≤ *V*(*j*) ≤ *r*, with *r* being the number of genes (12,849) from the drug profile. Then, *ES_up/down_* was set to *a_up/down_* if *b_up/down_*, and was set to -*b_up/down_* if *b_up/down_* > *a_up/down_*. Permutation tests repeated 100 times using randomly generated gene lists with the same number of up- and down-regulated genes as the HCoV signature data set were performed to measure the significance of the *ES* scores. Drugs were considered to have potential treatment effect if *ES* > 0 and *P* < 0.05, and the number of such HCoV signature data sets were used as the final GSEA score that ranges from 0 to 3. “NA” indicates that the drug cannot be evaluated by this method due to the lack of the drug profile.

## Data Availability

All codes and predicted repurposable drugs can be freely accessed at: https://github.com/ChengF-Lab/2019-nCoV

## AUTHOR CONTRIBUTIONS

F.C. conceived the study. Y.Z. and Y.H. developed the network methodology and performed all computational experiments. J.S., Y.Z., Y.H., Y.H., and W.M. performed data analysis. F.C., Y.Z. and Y.H. and J.S. wrote and critically revised the manuscript with contributions from other co-authors.

## Competing Interests

The content of this publication does not necessarily reflect the views of the Cleveland Clinic. The authors declare no competing interests.

## Notes

### Competing Interest Statement

The authors have declared no competing interest.

